# Cardiovascular drugs approved for heart failure with reduced ejection fraction and/or post-myocardial infarction exert consistent effects in both populations: A meta-analysis and meta-regression of randomized controlled trials

**DOI:** 10.1101/2024.02.05.24301181

**Authors:** Christopher Dasaro, Alyson Haslam, Vinay Prasad

**Author notes:** Corresponding author: Christopher Dasaro, BS, Lewis Katz School of Medicine at Temple University, 3500 N Broad St, Philadelphia, PA 19140. This author takes responsibility for all aspects of the reliability and freedom from bias of the data presented and their discussed interpretation.

## Abstract

**Background:** Heart failure (HF) following an acute myocardial infarction (post-MI HF) has been studied as an additional sub-type of HF to broaden the indications for HF drugs. Post-MI HF and HFrEF are pathophysiologically similar and share pharmacotherapies. In this meta-analysis, we examined the concordance between all-cause mortality data for drugs indicated for HFrEF and post-MI HF. We used our analysis to calculate the projected all-cause mortality hazard ratios (HRs) for the pending dapagliflozin (DAPA-MI) and empagliflozin (EMPACT-MI) post-MI HF trials.

**Methods:** Using CenterWatch and UpToDate, we identified all FDA-approved drugs for NYHA Class II to IV HFrEF. We searched each of these drugs on FDALabel and ClinicalTrials.gov to identify their registration trials measuring all-cause mortality for HFrEF and, if available, in the post-MI setting—including trials where participants displayed a left ventricular ejection fraction of <40% (“post-MI HF”). For each of the included studies, we extracted the all-cause mortality HRs, their 95% confidence intervals, and the control-group used. For all drugs studied in both indications, we plotted the all-cause mortality HRs for HFrEF against those for post-MI (HF) and calculated the linear regressions.

**Results:** This meta-regression pooled data from 29 completed trials underlying 20 drugs. Two pending trials were also analyzed. Nine drugs (metoprolol, carvedilol, spironolactone, eplerenone, sacubitril-valsartan, lisinopril, enalapril, valsartan, losartan) had all-cause mortality data in both HFrEF and post-MI generally, with a linear coefficient of determination of 0.93. Five of these drugs (carvedilol, eplerenone, sacubitril-valsartan, valsartan, losartan) were studied in both HFrEF and non-acute post-MI HF, displaying a linear coefficient of determination of 0.99. Using our model, we predict the all-cause mortality HRs that will be observed in the EMPACT-MI and DAPA-MI trials will be 0.85 and 0.89, respectively.

**Conclusions:** In this meta-regression of registration trials for drugs studied in both HFrEF and post-MI (HF), all-cause mortality effects were highly concordant. We also find asymmetries in the assessment of HF drug indications, whereby drugs are seldom assessed for an all-cause mortality benefit in both HFrEF and in post-MI HF. Future studies may use these results to guide future HF RCT development.

## Introduction

Heart Failure (HF) is a common clinical syndrome characterized by impaired cardiac output. HF can be characterized as having a preserved (HFpEF) or reduced (HFrEF) ejection fraction – with ≤40% defining the latter. In addition to HFpEF and HFrEF, heart failure following acute myocardial infarction (post-MI HF) has been studied as an additional sub-type of HF to broaden the indications for a given pharmacotherapy. This sequela is characterized by left ventricular dysfunction following an acute MI.

Myocardial infarctions are treated with several drugs–namely antiplatelet agents, beta blockers, statins, angiotensin-converting enzyme (ACE) inhibitors, angiotensin receptor blockers (ARBs), and mineralocorticoid receptor antagonists (MRAs) – with varied introduction of such therapies depending on the time since infarct^1^. Both HFrEF and post-MI HF are similar in pathophysiology and share medical therapies, yet both occupy unique niches for which treatments can claim market-share and broaden their indications.

We sought to examine the concordance of data supporting the use of select pharmacological therapies in HFrEF and post-MI HF. To do so, we aimed to characterize all of the regulatory trials underpinning the use of HFrEF agents in post-MI HF in terms of their all-cause mortality benefits (or lack thereof). In addition to clarifying the scope in which these drugs are indicated, our analysis provides grounds for preliminarily predicting outcomes in pending HF trials. In our case, we used this analysis to impute the projected all-cause mortality hazard ratios (HRs) for the pending dapagliflozin (DAPA-MI)^2^ and empagliflozin (EMPACT-MI)^3^ post-MI HF trials. We later broadened this model to predict the all-cause mortality HRs for drugs with missing trials for either HFrEF or post-MI HF.

## Methods

We systematically identified and characterized the trials supporting the approval of drugs indicated for HFrEF. First, we downloaded the UpToDate page detailing primary and secondary pharmacologic therapies for New York Heart Association (NYHA) functional classification II to IV HFrEF^4^. We also consulted CenterWatch^5^ for additional drugs labeled US Food and Drug Administration (FDA)-Approved for Heart Failure. All drugs included were confirmed to be approved by the US FDA.

From this list of US FDA-regulatory approvals for HF, we identified the registration clinical trials supporting their use for HFrEF and, if indicated, in the post-MI setting. For each drug, we searched on the FDALabel database^6^ and its associated Structured Product Labeling (SPL) document to determine the drug’s specific indication(s) and clinical trial(s) underpinning its approval. In instances where the SPL document failed to provide trial information (e.g trial name, NCT number), we searched for the drug’s registration trial on ClinicalTrials.gov using the particular drug as the “intervention”, and limited our search to randomized phase II, III, and IV trials in the English language. When reviewing the resulting trials, articles were included in our analysis only if all-cause mortality was a reported endpoint. We excluded articles that were pooled or secondary analyses; were retracted or inaccessible; contained fewer than 1,000 participants; were done in specific disease sub-populations (e.g only those with diabetes mellitus type 2); did not measure all-cause mortality, or only measured surrogate measures of morbidity (e.g., brain natriuretic peptide [BNP] levels).

We also characterized all of the post-MI studies by maximum LVEF permitted and the time between MI-onset and randomization. Noting the time-since-MI allowed us to assess how soon after the MI that a drug was initiated. Characterizing trials in this way may be relevant because the timing of LV dysfunction post-MI can logarithmically impact the risk of mortality^7^ and thus could obscure a direct comparison of the post-MI all-cause mortality benefits between drugs. A time-to-randomization of <3 days was considered the “acute” post-MI phase, and ≥3 days was considered the “non-acute” post-MI phase^7^.

For each of the included studies, we extracted the reported all-cause mortality HRs and their associated 95% confidence intervals. Additionally, we noted whether the trial used an active- or placebo-control; whether a trial existed for HFrEF, post-MI, or both; the number of participants randomized; and the LVEF and time-to-randomization used as inclusion criteria in any post-MI trials. For all drugs with both HFrEF and post-MI trials, we plotted the all-cause mortality HRs against each other in an x-y plane and calculated a linear regression. The confidence intervals, if available, for each condition are represented by the width and height of the ellipse surrounding that drug. All analyses were conducted using *Python*^8^.

We calculated regression coefficients and plotted the linear correlation for all drugs studied in HFrEF and post-MI– including both in the acute (time-to-randomization of <3 days) and non-acute (≥3 days) post-MI phase. We later isolated the drugs studied in HFrEF and the non-acute post-MI phase specifically, which incidentally isolated the post-MI trials where participants displayed a LVEF of ≤40% (“non-acute post-MI HF”). We also calculated and plotted separate correlations for drugs studied in either placebo- or actively-controlled trials.

From the resulting regression equations, we were able to (1) calculate the projected all-cause mortality HRs for the pending DAPA-MI and EMPACT-MI trials; and (2) estimate the all-cause mortality HRs for drugs with data in only one of the two indications. Specifically, we used this same model to impute the missing all-cause mortality metrics across multiple classes of drugs with missing data reported: ACE Inhibitors, ARBs, beta blockers, and MRAs/other class.

In accordance with 45 CFR §46.102(f), this study was not submitted for University of California, San Francisco institutional review board approval because it involved publicly available data and did not involve individual patient data.

## Results

Our search yielded 22 drugs from UpToDate and 11 from CenterWatch (**Table 1**). Eight drugs (sacubitril-valsartan, metoprolol, valsartan, dapagliflozin, empagliflozin, isosorbide/hydralazine, vericiguat, and ivabradine) were featured in both sources. In total, we found 23 unique drugs and 31 unique trials. Three drugs (ferric carboxymaltose, sotagliflozin, and canagliflozin) had registration trials done in specific disease sub-populations (e.g, exclusively those with iron-deficiency anemia or diabetes mellitus type 2), and were thus excluded from our analysis. Eight drugs had all-cause mortality data available for HFrEF only; three drugs had all-cause mortality data for post-MI only; and nine drugs (metoprolol, carvedilol, spironolactone, eplerenone, sacubitril-valsartan, lisinopril, enalapril, valsartan, losartan) had all-cause mortality data for both conditions (**Table 2**).

**Table 1:** Drugs included from UpToDate^A^ and CenterWatch^B^.

|  |  |
| --- | --- |
| Sacubitril-valsartan<sup>AB</sup> | Carvedilol<sup>A</sup> |
| Enalapril<sup>A</sup> | Bisoprolol<sup>A</sup> |
| Lisinopril<sup>A</sup> | Spironolactone<sup>A</sup> |
| Captopril<sup>A</sup> | Eplerenone<sup>A</sup> |
| Trandolapril<sup>A</sup> | Dapagliflozin<sup>AB</sup> |
| Ramipril<sup>A</sup> | Empagliflozin<sup>AB</sup> |
| Losartan<sup>A</sup> | Isosorbide/hydralazine<sup>AB</sup> |
| Candesartan<sup>A</sup> | Vericiguat<sup>AB</sup> |
| Valsartan<sup>AB</sup> | Ivabradine<sup>AB</sup> |
| Metoprolol<sup>AB</sup> | Digoxin<sup>A</sup> |

**Table 2:** Drug and trial characteristics for all HFrEF/post-MI therapies included in analysis.

|  | Number<br>(+pending**) |
| --- | --- |
| Number of included drugs from UpToDate and CenterWatch | 20 |
| Drugs with all-cause mortality data in either HFrEF or post-MI HF | 20 |
| Total number of trials included in analysis | 29 (2) |
| Drugs with all-cause mortality data for HFrEF, only | 9 (2) |
| Drugs with all-cause mortality data for post-MI, only | 3 |
| Drugs with all-cause mortality data for both HFrEF and post-MI | 9 (2) |
| Drugs with all-cause mortality data for both HFrEF and non-acute post-MI HF | 5 (2) |
| Drugs with all-cause mortality data for both HFrEF and post-MI HF; active-controls | 2 |
| Drugs with all-cause mortality data for both HFrEF and post-MI HF; placebo-controls | 4 (2) |
| Drugs with all-cause mortality data for both HFrEF and post-MI-HF, “mixed” active/placebo-controls | 2 |

\*\*Two trials, DAPA-MI and EMPACT-MI, are projected to be completed in June 2023 and August 2023, respectively

In addition to whether each drug was studied in both HFrEF and post-MI, we characterized the trials as having active-controls, placebo-controls, or a mix of the two. Of the nine drugs with all-cause mortality data in both conditions, five drugs (metoprolol, carvedilol, eplerenone, enalapril, and spironolactone) were studied against placebo in both indications. In contrast, two drugs (sacubitril-valsartan and losartan) were compared against active controls in both HFrEF and post-MI. Of note, only sacubitril-valsartan used two distinct active controls (enalapril in HFrEF; ramipril in post-MI); losartan was studied against captopril in both indications.

Two of the eight drugs (lisinopril and valsartan) had “mixed” active/placebo controls, whereby only one trial was placebo-controlled while the other had an active control. Specifically, lisinopril’s registration trial for HFrEF compared high (32.5-35 mg/day)- vs low (2.5mg-5mg)- dose lisinopril^9^, while its post-MI counterpart was placebo-controlled^10^. Valsartan, in contrast, was compared against placebo in HFrEF^11^ and against captopril in post-MI HF^12^.

Figure 1 shows the drugs’ all-cause mortality data reported in the post-MI survival trials as a function of those reported in HFrEF. The drugs are differentiated by control arm and have a gray ellipse representing the 95% confidence interval along either axis. Of note, the confidence intervals for metoprolol and enalapril were not provided in their registration trials for post-MI (MIAMI^13^ and CONSENSUS-II^14^, respectively) and thus do not have an ellipse. From this unadjusted regression model, we calculated an R^2^ of 0.87 and a correlation of 0.93.

**Figure 1:**
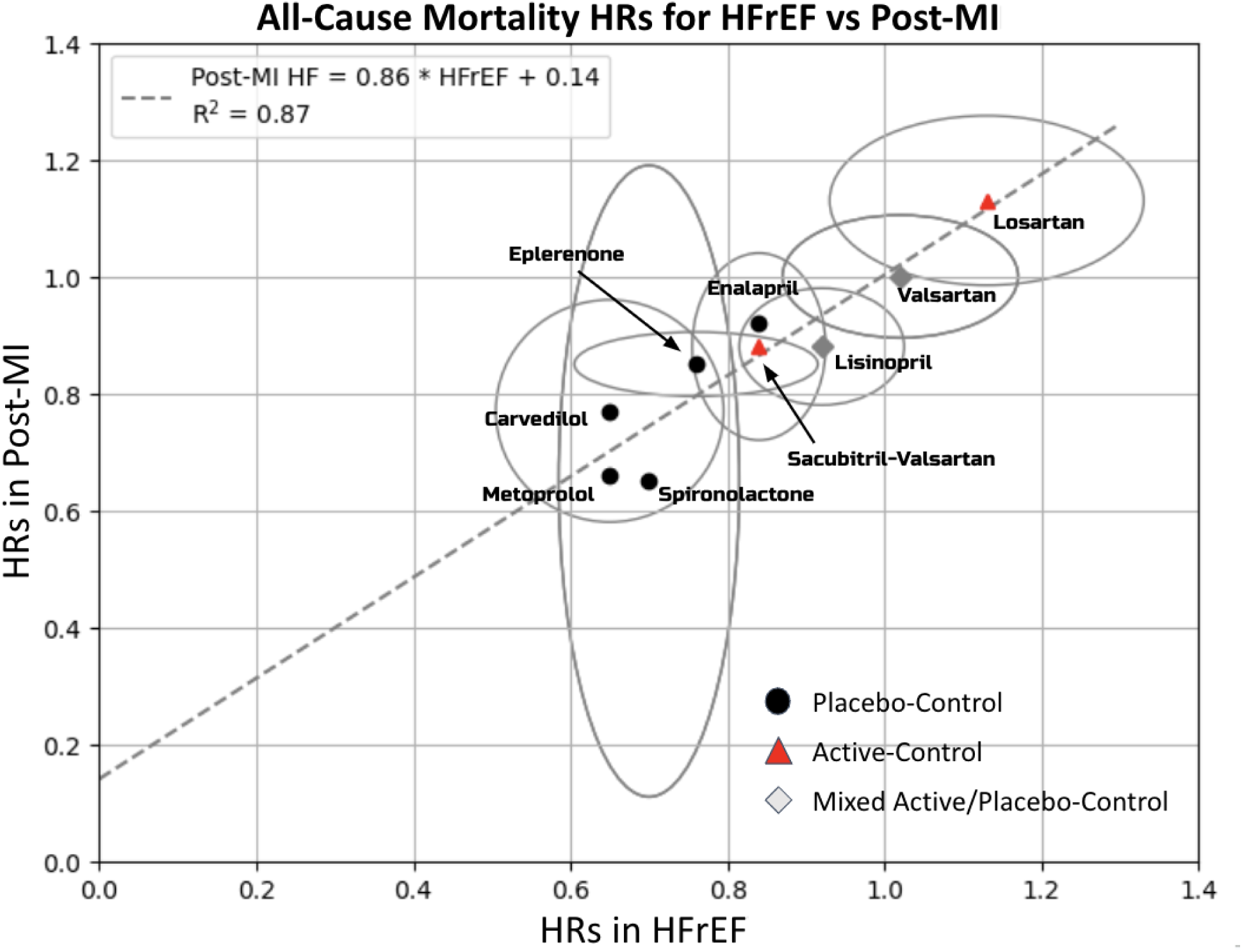
All-cause mortality data for drugs indicated for both HFrEF and post-MI. Drugs were labeled as being studied against placebo-controls, active-controls, or a combination (“mixed”). The height and width of the gray ellipses indicate the reported 95% confidence interval along either axis.

We further analyzed the post-MI trials to further characterize them by time-since-MI/time-to-randomization. We differentiated drugs that were studied in the acute post-MI phase (<3 days) from those studied in the non-acute (≥3 days) post-MI phase. Figure 2 illustrates the time-since-MI inclusion criteria, in days, for all of the post-MI trials included in our analysis that reported such data. Four trials (GISSI-3, ALBATROSS^15^, CONSENSUS-II, and MIAMI) studied their respective drug (lisinopril, spironolactone, enalapril, and metoprolol) only in the acute post-MI phase. Past reports indicate that the development of HF more than 3 days post-MI is associated with a 43% increase in mortality than when developed in the first 3 days^7^. Thus, we re-analyzed the data in Figure 1 after removing these trials, resulting in an improvement in the coefficient of determination (R^2^ of 0.98, Figure 3). Excluding these trials also allowed us to isolate all of the post-MI trials that only allowed LVEFs ≤40% (“post-MI HF”) (**Table S1**).

**Figure 2:**
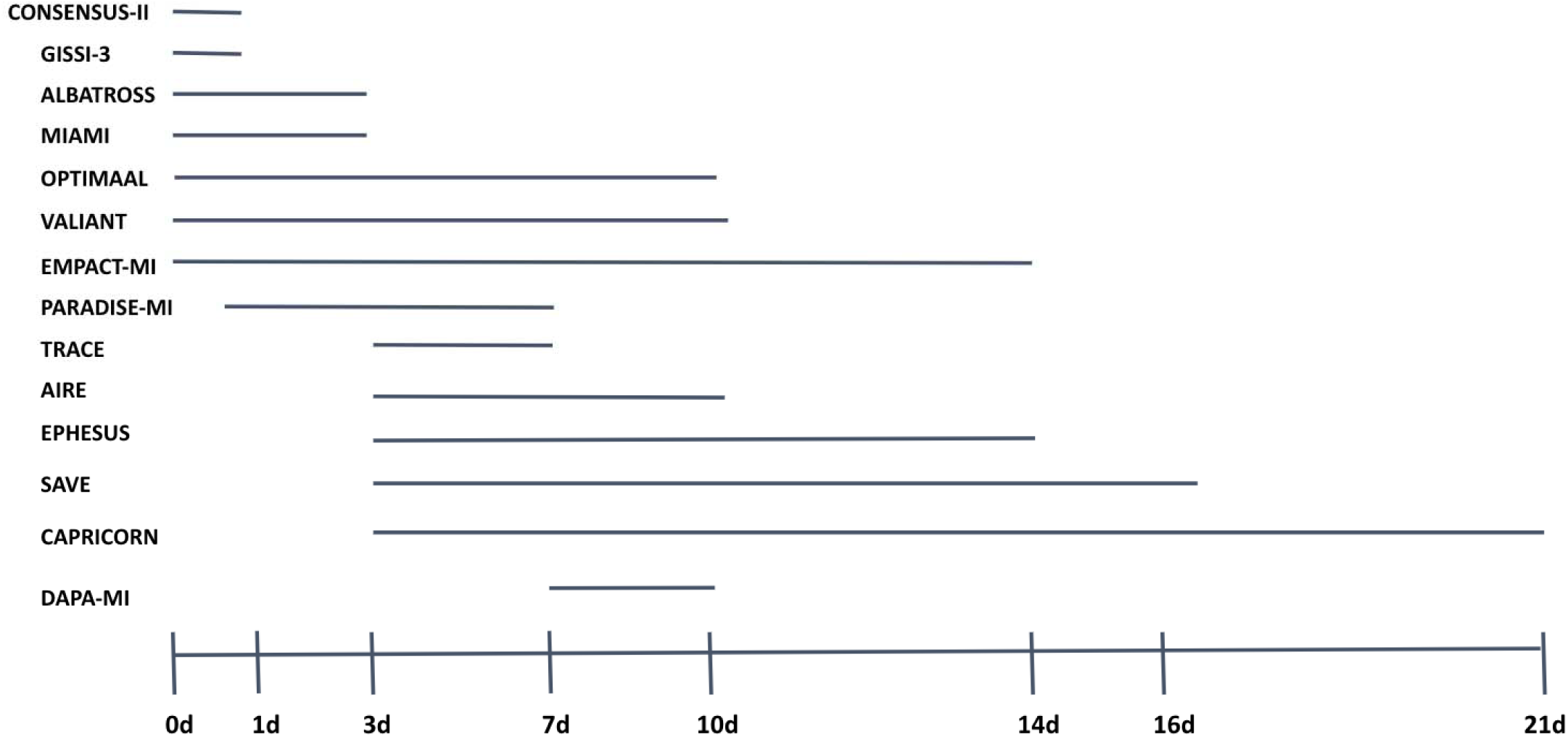
Time between onset of MI and randomization, reported in days, in post-MI trials.

**Figure 3:**
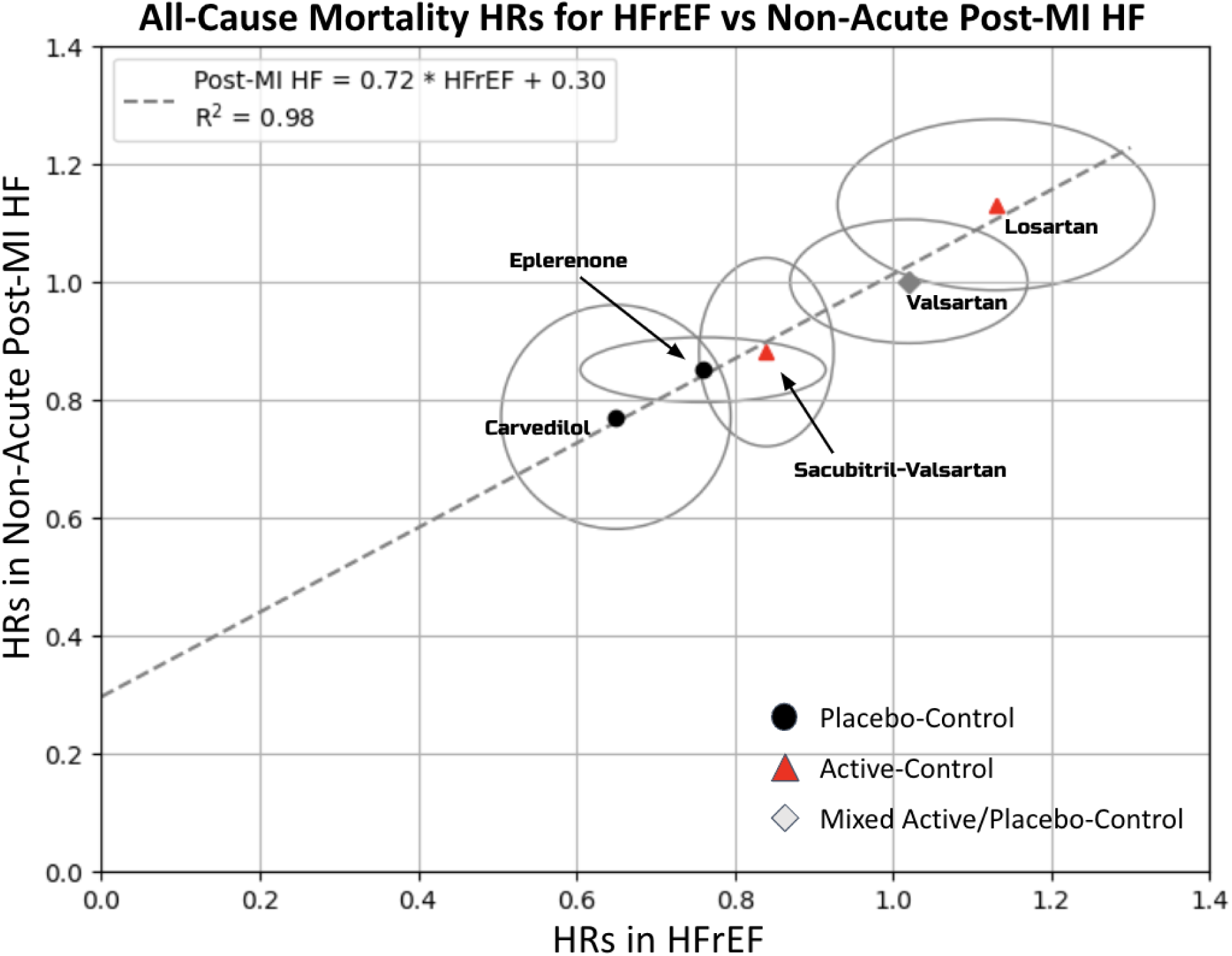
All-cause mortality data for drugs indicated for both HFrEF and non-acute (>3 days) post-MI heart failure.

Figure 4A and **4B** splits drugs tested against placebo (in at least one of its trials) and those tested against an active-control (in at least one of its trials). We found coefficients of determination of 0.98 among the active-controlled trials and 1.00 in the placebo-controlled trials. As EMPACT-MI and DAPA-MI are projected to be placebo-control trials, we imputed their all-cause mortality benefits from the linear regression calculated in Figure 4A. This model predicted all-cause mortality HRs of 0.89 and 0.85, for trials of DAPA-MI and EMPACT-MI, respectively (Figure 4A).

**Figure 4A:**
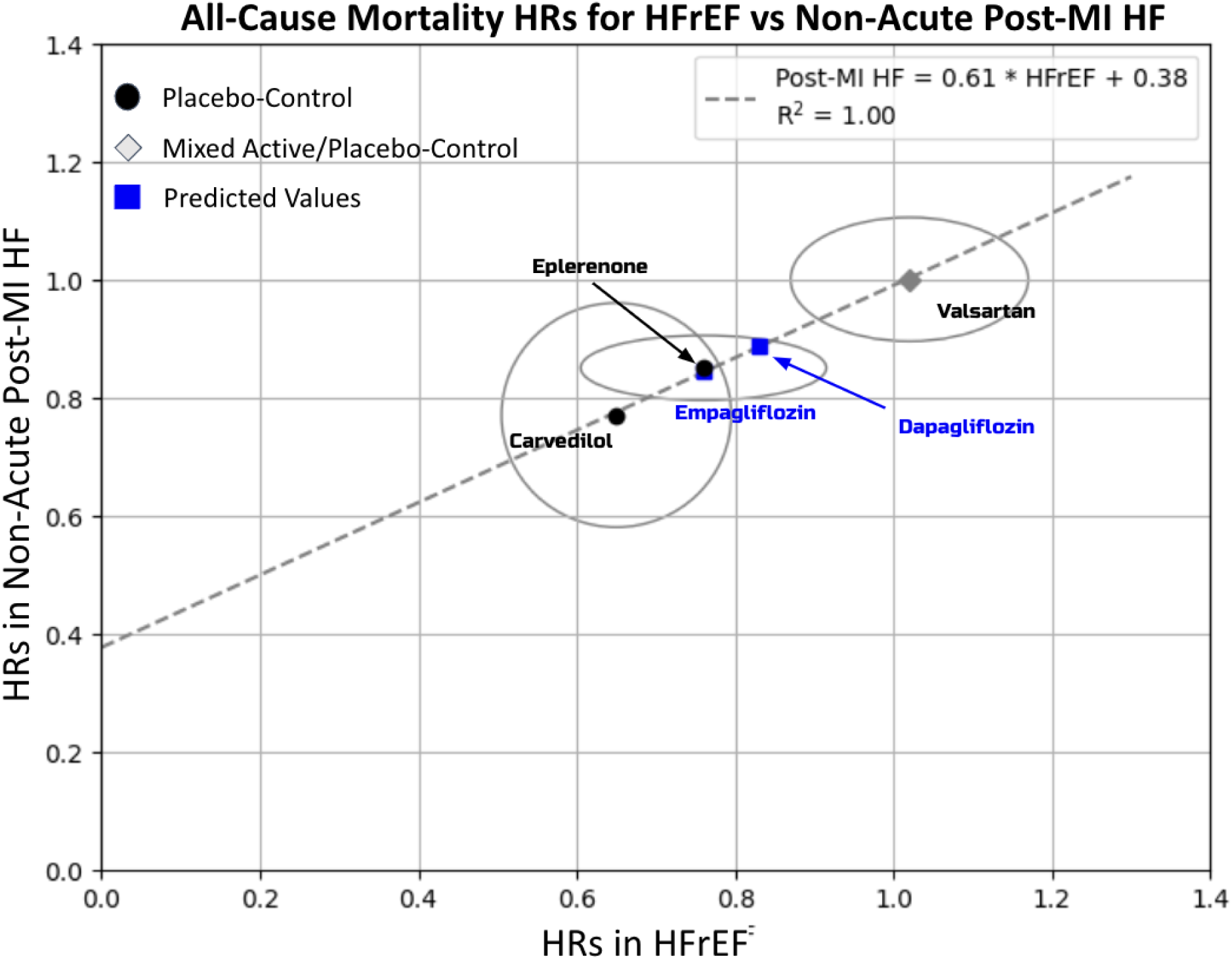
Placebo-controlled HF drugs with EMPACT-MI (empagliflozin) and DAPA-MI (dapagliflozin) HR predictions of 0.85 and 0.89, respectively.

**Figure 4B:**
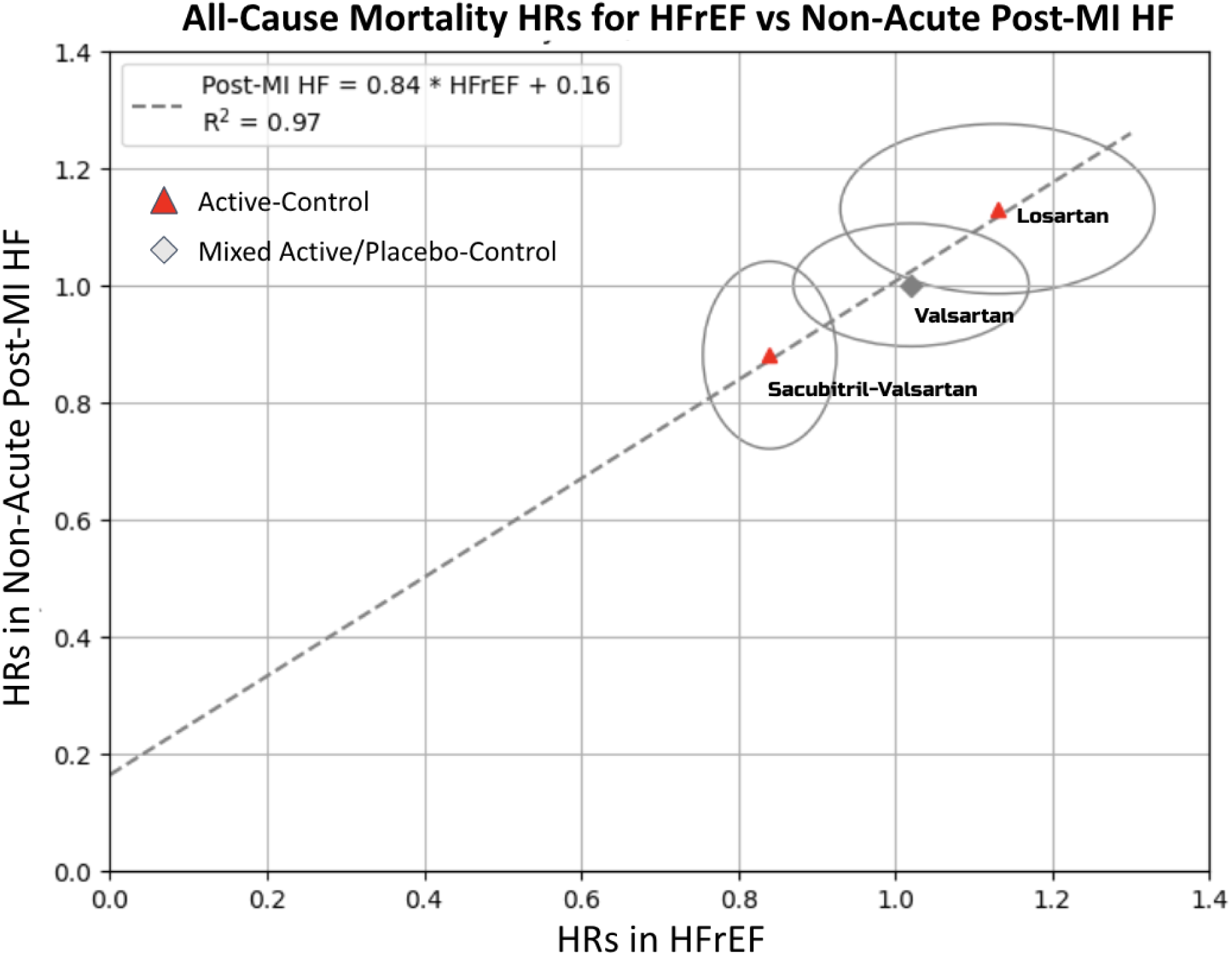
Active-controlled drugs indicated for HFrEF and post-MI HF.

Using the placebo-controlled linear regression modeled in Figure 4A, we repeated our all-cause mortality predictions for all drugs that had missing all-cause mortality data in either HFrEF or post-MI HF. We broke up this analysis by drug class and reported the HRs as cartesian coordinates (**Figure S1, panels A-D**). Figure S1 shows the missing all-cause mortality metrics across multiple classes of drugs with missing data reported: ACE Inhibitors (panel A), ARBs (panel B), MRAs/other class (panel C), and beta blockers (panel D).

## Discussion

We found among 23 unique drugs indicated for HF, only nine (39%) currently have trials measuring all-cause mortality in both NYHA II-IV HFrEF and post-MI. When accounting for drugs that were studied only in the acute phase following an MI and limited patients to a LVEF of <40%, only five (22%) drugs measured all-cause mortality in both HFrEF and non-acute post-MI HF. Of these five drugs, two were studied in placebo-controlled trials while three partially or entirely used active-controls. Conversely, eight drugs (35%) had survival data for only HFrEF, while three (13%) others had data only for post-MI HF. These data points to pervasive asymmetries in the treatment of HF, whereby drugs are seldom assessed for an all-cause mortality benefit in both HFrEF and in post-MI HF.

Using the model in Figure 4A, we calculated the expected all-cause mortality HRs for the pending dapagliflozin and empagliflozin post-MI trials. Per the published protocols, the EMPACT-MI and DAPA-MI are placebo-controlled trials. We estimate the HR in these ongoing trials to be 0.85 and 0.89, respectively.

Similarly, we used this strategy to extrapolate the all-cause mortality HRs for 12 (52%) drugs missing either HFrEF or post-MI HF data (Figure S1A-D). We grouped this analysis by drug class: ACE inhibitors, ARBs, beta blockers, and MRAs/other classes. The ACE inhibitor class had the most “missing” data, with this analysis filling in all-cause mortality gaps for four drugs. This is because the efficacies of three ACE inhibitors–captopril, trandolapril, and ramipril–were studied in three placebo-controlled post-MI HF trials (SAVE (1992)^16^, TRACE (1995)^17^, and AIRE (1993)^18^, respectively), but not in HFrEF. The extrapolation of all-cause mortality data in post-MI HF to HFrEF (though in the absence of confidence intervals) shows plausible all-cause mortality benefits for these drugs. A possible reason for such strong benefits could be that most patients in these studies were treated with fibrinolytic therapy or no reperfusion; data in patients who underwent percutaneous coronary intervention (PCI) post-MI are limited^19^.

Though excluded from the subsequent post-MI analysis shown in Figure 3, the ALBATROSS trial technically did not study the addition of spironolactone vs. placebo. Instead, its intervention was the addition of an MRA regimen of potassium canrenoate bolus followed by 6 months of oral spironolactone. As a result, direct comparisons between this trial and RALES^20^ (HFrEF trial) are difficult to make. Furthermore, the confidence intervals reported for all-cause mortality in ALBATROSS were uncharacteristically large in our figure. This could be because the all-cause death analysis was done in a non-pre-specified exploratory fashion. If done at an early time point in the study, this could explain the large confidence intervals and could lead to type I errors.

Figure 3 suggests that sacubitril-valsartan is the only drug that reportedly provides an all-cause mortality benefit in HFrEF, but not post-MI HF. Prior work^21^ has noted that sacubitril-valsartan was studied in a fairly unique [A + B] vs. C design ([sacubitril + valsartan] vs. enalapril) – a format that continued in its post-MI study PARADISE-MI^22^ ([sacubitril + valsartan] vs. ramipril). Taken together, that the survival trials for sacubitril-valsartan are unique in both design and outcomes may suggest the need for further trials^32^. Possible trial designs–one that adopts an [A + B] vs. placebo or [A + B] vs. B design–already exist for sacubitril-valsartan in HFpEF^24^ and in an analysis of NYHA IV HFrEF^25^. Notably, the former study was negative for both combined heart failure hospitalizations/cardiovascular mortality and all-cause mortality, and the latter was negative for both primary and secondary endpoints (none of which were all-cause or cause-specific mortality).

Figure 1 suggests that lisinopril shows similarly unique results–demonstrating an all-cause mortality benefit in post-MI but not HFrEF; however, its registration trial for HFrEF, ATLAS, reports a combined all-cause morbidity-mortality benefit in HFrEF. This comparison is further complicated by the fact that ATLAS compared two doses of lisinopril, whereas sacubitril-valsartan was compared against ramipril. Additionally, as Figure 3 excludes lisinopril’s post-MI trial (GISSI-3), sacubitril-valsartan appears truly unique among non-acute post-MI HF trials in demonstrating an asymmetric all-cause mortality benefit. Figure 2 shows that the inclusion criteria for PARADISE-MI allowed for participants whose MI was between 0.5 and 7 days prior to randomization. This trial therefore did allow for some acutely post-MI participants; however, the distribution of time-to-randomization was not reported.

## Strengths/Limitations

The major strength of this study is that this is the first comprehensive analysis of all drugs indicated for heart failure of its kind, and highlights numerous gaps in our understanding of these drugs’ benefits and studied indications. We relied on two data sources (UpToDate and CenterWatch) to generate a comprehensive list of FDA regulatory approvals for heart failure.

There are four limitations to our study. (1) Our search criteria encompass only trials that measured all-cause mortality as a primary or secondary endpoint. This can be considered a limitation in situations where other metrics of morbidity or mortality are assessed, but not all-cause mortality. Additionally, given the diversity of trial designs studied here, a direct head-to-head comparison between any two drugs can be muddied by the nuances within any given trial– e.g, the presence or absence of a run-in period, the dose of each drug used, etc.

(2) Our search criteria yielded nine drugs in our initial analysis and five drugs in the subsequent analysis. Given the limited number of drugs available for analysis, the true strength of the linear relationship established in our regression model may be tenuous or uncertain; future trials may provide more data points to modulate the relationship.

(3) Though our analysis of post-MI trials focused on the times-to-randomization and LVEF, there were other sub-analyses that could have been done for this set of trials. Many trials report sub-analyses divided by location of myocardial infarct (e.g anterior, anterolateral) or the presence/absence of ST-elevations on electrocardiogram. As another example, the SAVE and AIRE trials–though both post-MI HF trials–differed in whether the patients were in symptomatic HF despite both trials including only patients with an ejection fraction of <40%. These groupings are certainly additional opportunities for further characterizing post-MI trials that could provide insight into contexts for which particular drugs are more or less efficacious.

(4) This analysis was restricted to only registration survival data and included no long-term follow-up metrics of morbidity or mortality. Thus, any long-term reports that further guided or modulated the use of any drugs listed here–for example, a follow-up analysis reporting all-cause mortality benefit despite the registration trial not reporting such–is not accounted for in this analysis. Similarly, our extrapolation of incomplete data assumes a linear relationship between HFrEF and post-MI HF and did not allow us to impute confidence intervals for any HR predictions.

## Conclusion

Our study provides important insights into the current asymmetries in assessing the indications for heart failure drugs. We found that only five of 23 drugs indicated for heart failure have been studied in both HFrEF and post-MI HF. Our imputation of all-cause mortality metrics for dapagliflozin, empagliflozin, and other drugs may provide insight into future directions for clinical trials of currently approved heart failure drugs. We predict that the trials DAPA-MI and EMPACT-MI will report HRs of 0.89 and 0.85, respectively. As these two studies come to completion in the summer of 2023, our modeling will be further modulated and possibly substantiated. Older therapies that lack randomized trials may also be tested by non-conflicted bodies, including the VA or National Institutes of Health.

## Data Availability

All data produced in the present study are available upon reasonable request to the authors

## Contributors

VP conceptualized study design; CD reviewed the literature; CD curated data; VP and AH reviewed and confirmed abstracted data; CD wrote the first draft of the manuscript and all authors reviewed and revised subsequent and finalized draft of the manuscript

---

**Figure S1A-D:**
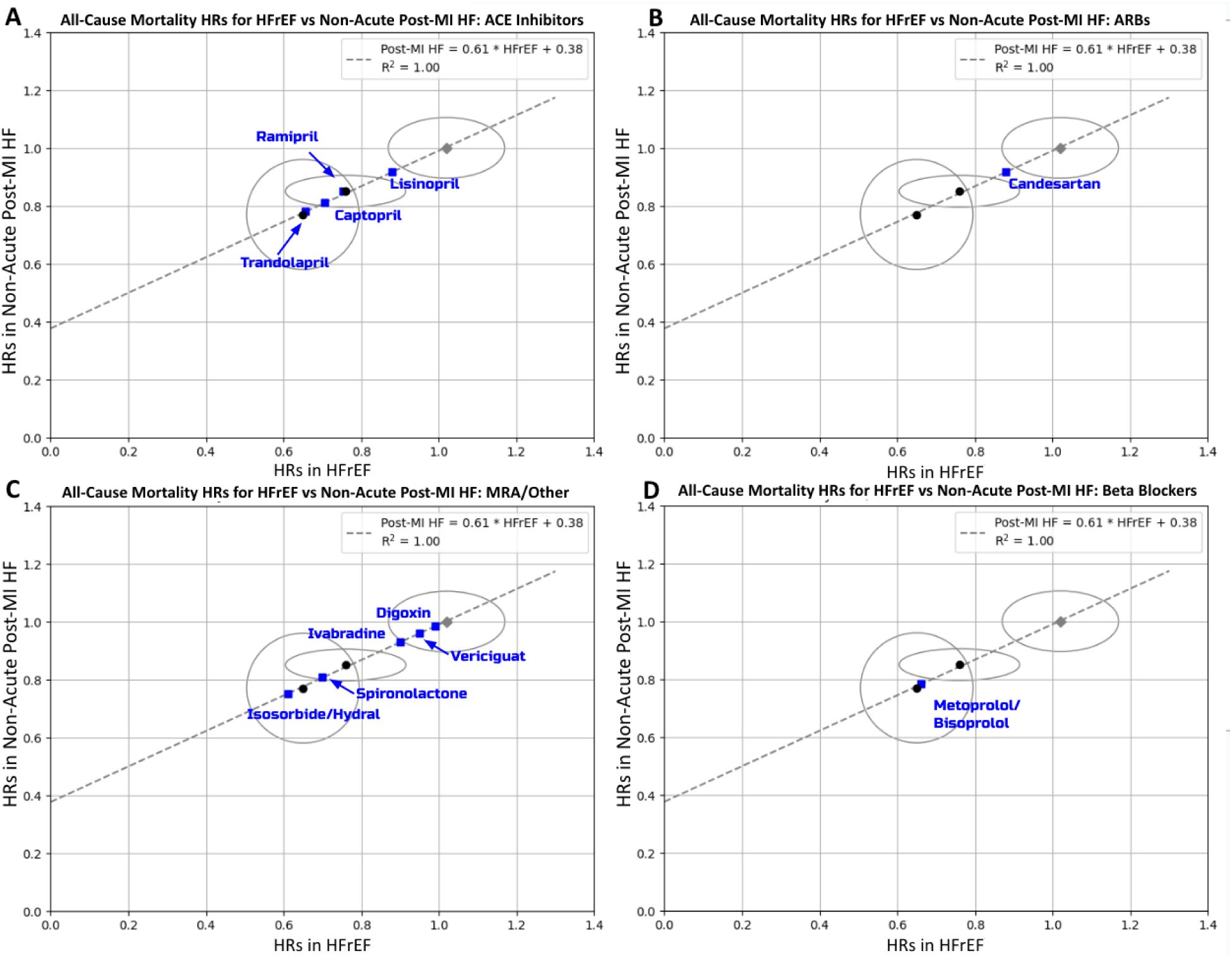
Predicted all-cause mortality metrics for 12drugs that were missing either HFrEF or post-MI HF data. The linear regression to which they were fitted is the same as the placebo-controlled regression. These drugs are broken up by class with all-cause mortality metrics given as cartesian coordinates (HFrEF, post-MI HF); * indicates predicted value from regression: **A)** ACE inhibitors: trandolapril (0.66*, 0.78); captopril (0.70*, 0.81); ramipril (0.75*, 0.84); lisinopril (0.92, 0.88*). **B)** ARBs: candesartan (0.88, 0.92*). **C)** MRAs and other classes: isosorbide/hydralazine (0.61, 0.75*); spironolactone (0.70, 0.81*); ivabradine (0.90, 0.93*); vericiguat (0.95, 0.96*); digoxin (0.99, 0.98*). **D)** Beta blockers: metoprolol (0.66, 0.78*); bisoprolol (0.66, 0.78*).

**Table S1:** Characteristics of post-MI trials.

| Post-MI Trial | LVEF (%) | Participants Randomized | Intervention vs Control | All-Cause Mortality HR/RR (95% CI) |
| --- | --- | --- | --- | --- |
| MIAMI (1985) | N/a | 5,778 | Metoprolol vs Placebo | 0.64 (N/a) |
| SAVE (1992) | <40% | 2,231 | Captopril vs Placebo | 0.81 (0.68-0.97) |
| CONSENSUS-II | N/a | 6,090 | Enalapril vs Placebo | 0.92 (N/a) |
| AIRE (1993) | <40% | 2,006 | Ramipril vs Placebo | 0.84 (0.75-0.95) |
| GISSI-3 (1994) | N/a | 19,394 | Lisinopril vs Placebo | 0.88 (0.79-0.99) |
| TRACE (1995) | <35% | 1,749 | Trandolapril vs Placebo | 0.78 (0.67-0.91) |
| CAPRICORN (2001) | <40% | 1,959 | Carvedilol vs Placebo | 0.77 (0.60-0.98) |
| OPTIMAAL (2002) | <35% | 5,477 | Losartan vs Captopril | 1.13 (0.99-1.28) |
| VALIANT (2003) | <35% | 9,818 | Valsartan vs Captopril | 1.00 (0.90-1.10) |
| EPHESUS (2003) | <40% | 6,642 | Eplerenone vs Placebo | 0.85 (0.75-0.86) |
| ALBATROSS (2016) | 50% ** | 1,616 | MRA regimen vs Std of Care | 0.65 (0.30-1.38) |
| PARADISE-MI (2021) | <40% | 5,211 | Sacubitril-valsartan vs Ramipril | 0.88 (0.73-1.05) |
| DAPA-MI (2023) | <40% | 4,017 | Dapagliflozin vs Placebo | TBA |
| EMPACT-MI (2023) | <45% | 6,522 | Empagliflozin vs Placebo | TBA |

\*\*The reported LVEF range was 45-60%; the average in both arms was 50%

**Table S2:** Characteristics of HFrEF trials.

| **HFrEF Trial** | **# Randomized** | **Intervention vs Control** | **All-Cause Mortality HR/RR (95% CI)** |
| --- | --- | --- | --- |
| SOLVD (1991) | 2,549 | Enalapril vs. placebo | 0.84 (0.74-0.95) |
| DIG (1997) | 6,800 | Digoxin vs placebo | 0.99 (0.91-1.07) |
| ATLAS (1999) | 3,164 | High vs low dose lisinopril | 0.92 (0.82-1.03) |
| CIBIS II (1999) | 2,647 | Bisoprolol vs placebo | 0.66 (0.54-0.81) |
| MERIT-HF (1999) | 3,991 | Metoprolol vs placebo | 0.66 (0.53-0.81) |
| RALES (1999) | 1,663 | Spironolactone vs placebo | 0.70 (0.69-0.81) |
| ELITE II (2000) | 3,152 | Losartan vs captopril | 1.13 (0.97-1.35) |
| COPERNICUS (2001) | 2,289 | Carvedilol vs placebo | 0.65 (0.52-0.81) |
| Val-HeFT (2001) | 2,289 | Valsartan vs placebo | 1.02 (0.88-1.18) |
| A-Heft (2004) | 1,050 | Isosorbide/hydralazine vs placebo | 0.61 (N/a)* |
| CHARM (2004) | 4,576 | Candesartan vs placebo | 0.88 (0.79-0.97) |
| SHIFT (2010) | 6,558 | Ivabradine vs placebo | 0.90 (0.80-1.02) |
| EMPHASIS-HF (2011) | 2,737 | Eplerenone vs placebo | 0.76 (0.62-0.93) |
| PARADIGM-HF | 8,399 | Sacubitril-valsartan vs enalapril | 0.84 (0.76-0.93) |
| DAPA-HF (2019) | 4,744 | Dapagliflozin vs placebo | 0.83 (0.71-0.97) |
| EMPEROR-Reduced (2020) | 3,730 | Empagliflozin vs placebo | 0.76 (0.62-0.93) |
| VICTORIA (2020) | 2,526 | Vericiguat vs placebo | 0.95 (0.85-1.07) |

